# Epidemiology of Autism Spectrum Disorder in Buenos Aires, Argentina: Insights from Unique Disability ID Issuance Trends

**DOI:** 10.1101/2023.07.08.23292383

**Authors:** Agustina Aragón-Daud, Giselle Vetere, Marina Argañaraz, Francisco Musich

**Author notes:** Corresponding author Address: Marcelo T. de Alvear 1632, Buenos Aires, Argentina.

## Abstract

**Purpose:** Autism Spectrum Disorder (ASD) prevalence rates vary greatly across regions and studies. Some countries, such as Argentina, have unknown rates. Due to this high heterogeneity in ASD prevalence worldwide, it is important to study its prevalence and characteristics in such countries to develop effective policies.

**Methods:** we analyzed official data from the Unique Disability IDs (UDIDs) issued for individuals with ASD in the Autonomous City of Buenos Aires from 2016 to 2021, examining age, gender, and diagnosis.

**Results:** most UDIDs were issued for males, for Pervasive Developmental Disorders, and almost half to individuals over 8 years old, with these trends remaining stable over these years. However, UDID issuance abruptly dropped in 2020.

**Conclusions:** our findings highlight epidemiological aspects of the autism population in Argentina, including challenges such as delayed diagnosis and high prevalence of broad diagnosis categories. Addressing these challenges requires further research and intervention to improve the quality of life of individuals with ASD.

---

To be inclusive of diverse voices within the community (Bottema-Beutel et al., 2021; Vivanti, 2020), in this study, we will use both person-first (“person with autism”) and identity-first (“autistic”) language interchangeably when referring to autism. We believe that this flexible approach to terminology better reflects the dynamic nature of both research and the autistic community.

Autism is a neurodevelopmental condition that affects communication, social interaction, and behavior (World Health Organization [WHO], 2023). It is a highly prevalent mental disorder, yet the prevalence figures for Autism Spectrum Disorder (ASD) are quite heterogeneous. For example, while the WHO (2023) estimates that 1 in 100 children has ASD, the Centers for Disease Control and Prevention (CDC, 2021) estimates that 1 in 44 children is diagnosed with some form of ASD. Many factors are thought to account for these differences in prevalence, such as the population sampled, the study design, and the diagnostic criteria used (Elsabbagh et al., 2012; Talantseva et al., 2023).

The prevalence of ASD varies widely across different regions. According to a review of 71 studies conducted in the United States and different countries in Europe between 2012 and 2021, the prevalence of ASD ranged from 1 to 436 per 10.000 people (Zeidan et al., 2022). In Latin America, however, the estimated prevalence was between 25 and 30 per 10.000 people, based on data from 2011 to 2013 (Fajardo et al. 2021). Additionally, in many low- and middle-income countries, such as Argentina, the prevalence of ASD is largely unknown (Elsabbagh et al., 2012; Valdez et al., 2021; WHO, 2023), which can hinder the development of effective policies to support individuals with ASD and their families. Therefore, it is imperative to obtain further epidemiological data in these countries to adequately address this matter.

Moreover, several studies suggest that the prevalence of autism is increasing (CDC, 2021; Elsabbagh et al., 2012; Fajardo et al., 2021; Talantseva et al., 2023). The explanations for this phenomenon are diverse, ranging from expansions in diagnostic criteria (such as increased diagnoses of ASD and decreased diagnoses of Intellectual disability), environmental factors (such as increased survival rates of premature babies, who have a higher incidence of ASD, as found by Wang et al., 2017), advanced parental age (as reported by Kim et al., 2019), and increased awareness of the disorder and access to healthcare services (APA, 2022).

Despite this considerable variability in autism epidemiology, there are some well-established aspects. First, one of the most notable differences is that autism is more prevalent among males than females (APA, 2022; Elsabbagh et al., 2012; Hsu et al., 2012). Biological and genetic evidence supports this observation (Bölte et al., 2023). However, other variables may also contribute to this difference in prevalence. For instance, females with autism often exhibit camouflaging, which is the ability to mask autistic symptoms and can result in under diagnosis (Hull et al., 2020; Kirkovski et al., 2013; Loomes et al., 2017).

Second, signs of autism, as a neurodevelopmental disorder, can manifest in children as early as 12 to 24 months of age (APA, 2022). However, prevalence rates are higher among older children, aged 6 to 12 years, than those under 5 years old (Talantseva et al., 2023). This trend is exemplified in a study conducted in Latin American and Caribbean countries, where concerns about developmental delays appeared before the age of 2 but the average age at diagnosis was 3-4 years old (Montiel-Nava et al., 2023). This is concerning, as early detection and communication of ASD is associated with a better prognosis (Freitas & Revoredo, 2022; Molina, 2012; Webb & Jones, 2017) and increased self-understanding and empowerment in the patient (Oredipe et al., 2022).

Autism is a spectrum disorder, meaning that there is a wide range of abilities and impairments that occur among those with the diagnosis. While some autistic people may have average or above-average intelligence and need little support to function independently, approximately 75% of individuals with autism have moderate to severe intellectual disability, limited verbal communication, and limited adaptive behavior (APA, 2022). Due to this, autism is considered a cause of disability in many countries, including Argentina. Autistic people in Argentina, as well as their caregivers, can request a Unique Disability ID (UDID) voluntarily.

The UDID is a public document in Argentina valid throughout the country and guarantees a variety of rights, such as 100% coverage for disability-related diagnostic procedures, family allowances, transportation, and International Symbol of Accessibility (ISA). It is issued by a specialized team of healthcare professionals after a thorough interdisciplinary evaluation of the person’s developmental history, symptoms, and functioning. The official nomenclature used for issuing UDIDs for autism in Argentina is the International Classification of Diseases (ICD-10) of the WHO (1992). According to the ICD-10, ASD falls under the category of Pervasive Developmental Disorders (PDD, F84).

Furthermore, inside the PDD category, Argentina considers Infantile autism (F84.0), Atypical autism (F84.1), Asperger’s syndrome (F84.5), other PDD (F84.8), and PDD not otherwise specified (PDD-NOS, F84.9). Lastly, as of March 2023, UDIDs no longer expire and therefore no longer require renewal.

Although there has been an increase in requests for family guidance related to autism in the Autonomous City of Buenos Aires (Argentina), indicating a growing awareness of ASD in the country (Puga et al., 2019), there is currently a lack of epidemiological data on autism in Argentina and its local regions. Moreover, the high heterogeneity of worldwide prevalence rates on autism makes it difficult to extrapolate them to the Argentinian population. Numerous studies have emphasized the urgent need for further research in low- and middle-income countries (Elsabbagh et al., 2012; Durkin et al., 2015; Valdez et al., 2021; WHO, 2023). Studying the issuance of UDIDs for autism in the Autonomous City of Buenos

Aires could provide an estimate of the incidence of autism and its sociodemographic characteristics. In this study, we examine the trends and patterns of autism UDID issuance over time, explore the demographic factors associated with autism, and discuss the implications of our findings for autism research and policy in Argentina.

## Methods

This retrospective descriptive study examines the issuance of UDID numbers in the Autonomous City of Buenos Aires, Argentina, from 2016 to 2021 for individuals with ASD. The data was obtained from the Disability Assessment Board of the government of the city. These official data are difficult to access as they are not publicly available and require authorization. Demographic characteristics, including gender and age, were analyzed alongside diagnoses within the ASD spectrum according to the ICD-10. While the data were limited to descriptive analysis, this study is the first of its kind in the country. We used R software (version 4.2.2) to perform the data analysis.

## Results

### General trends in the issuance of UDIDs for autism

Between 2016 and 2021, 6621 UDIDs were issued due to autism in the Autonomous City of Buenos Aires, Argentina. Of these UDIDs, 80% (*n* = 5303) remain active, while 20% (*n* = 1318) had expired. More than half of them have been issued to males (63.54%, *n* = 4207), while the remaining UDIDs were granted to females. Roughly half have been granted for children aged 1-7 years old (54.87%, *n =* 3633). In this way, the reminding 45.13% (*n* = 2988) have been issued for older ages: children aged 8-18 accounted for 38.54% (*n =* 2552) of the UDIDs, while adults represented 6.59% (n = 436) of the UDIDs.

Following the ICD-10 classification, the most prevalent diagnosis by far was PDD, accounting for 43.21% (*n* = 2861) of the cases. The remaining diagnoses were evenly distributed: Infantile Autism (15.24%, *n* = 1009), PPD-NOS (11.78%, *n* = 780), other PDD (10.48%, *n* = 694), Asperger (10.16%, *n* = 673), and Atypical Autism (9.12%, *n* = 604).

### Trends by year in the issuance of UDIDs for autism

According to Fig. 1a, the largest number of UDIDs was issued in 2017, with a total of 1545. However, since 2020, there has been a significant decline in the number of UDIDs issued. In fact, in each year since 2020, less than 800 UDIDs have been issued, which is a considerable drop compared to the consistently high number of issuances exceeding 1100 in previous years.

**Figure 1.**
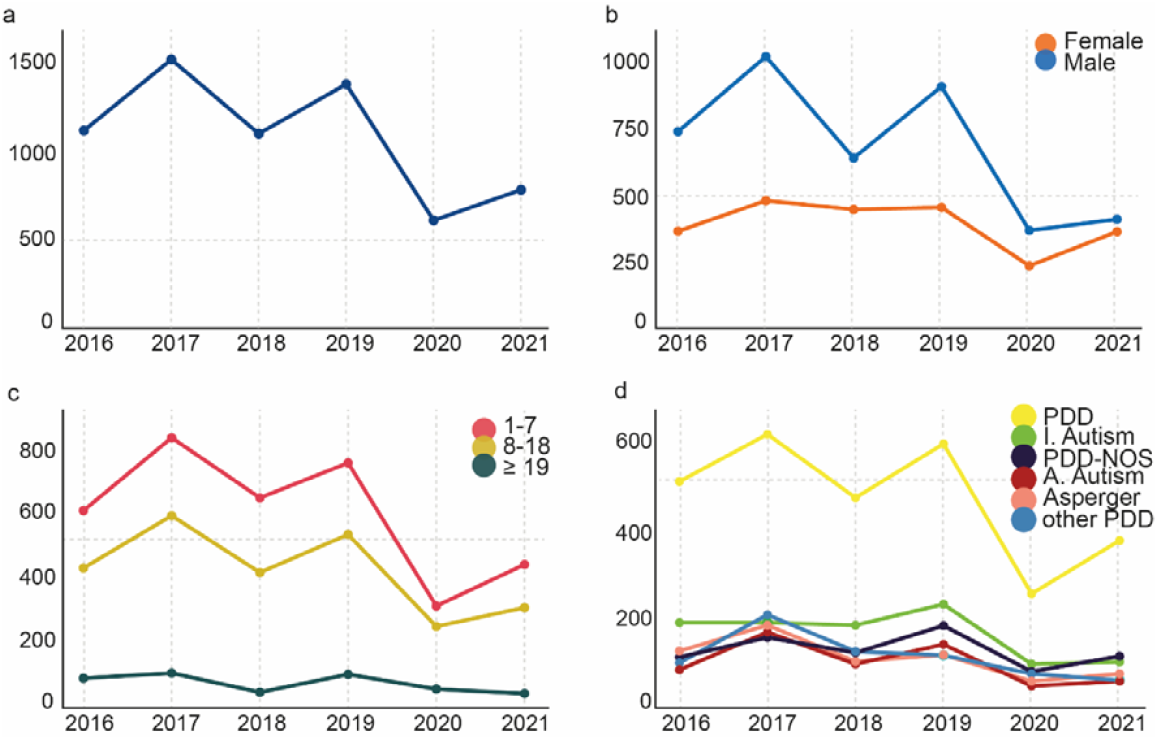
Absolute frequency of issuance of UDIDs per year in total (a) and according to gender (b), age (c), and diagnoses (d).

Regarding gender, every year from 2016 to 2021, the number of UDIDs issued to males was consistently higher than that issued to females, around 20% higher in most years. However, this difference has gotten smaller in the last two years (Fig. 1b), with only a difference of 6% in the last year.

Regarding age, every year the amount of UDIDs issued has been larger for children aged 1-7 years old, followed by children aged 8-18 years old, and lastly, by adults (Fig. 1c). In the last year, there has been a slight decrease in the number of UDIDs issued for adults, accounting only for 5%, while the UDIDs issued for children between 8 and 18 years old remains quite high, accounting for 39% of them.

Regarding diagnosis, PDD was the most frequently diagnosed condition for the issuance of UDIDs every year. Infantile Autism was the second most commonly diagnosed condition, although in 2017 there have been more UDIDs issued for other types of PDD, and in 2021, there were more UDIDs issued for PDD-NOSS. In terms of the other diagnoses, the number of UDIDs issued was similar among them (Fig 1d). Notably, the issuance of UDIDs for initial diagnoses was consistently higher than the issuance for renewals across all conditions each year.

## Discussion

This is the first study to explore official registers of UDIDs for autism in Argentina, focusing on the Autonomous City of Buenos Aires. We analyzed general trends from 2016 to 2021 and found that UDID issuance is higher for males, with PDD being the most prevalent diagnosis, and almost half of them issued for individuals over 8 years old. Despite the number of UDIDs issued decreasing abruptly in 2020, these trends persist every year, with some minor fluctuations that we discuss below.

### General trends in the issuance of UDIDs for autism

Between 2016 and 2021, a total of 6621 individuals obtained their UDIDs for autism.

The majority of these individuals were male, which is in line with the higher prevalence of ASD in males compared to females. The worldwide prevalence of ASD in males is typically reported to be four times higher than in females (APA, 2022), although some studies suggest it may be closer to three times higher (Loomes et al., 2017). This gender difference in ASD prevalence may be partly due to female-specific characteristics such as camouflaging, which can mask ASD symptoms in females (Hervás, 2022; Lai et al., 2017; Ruggieri & Arberas, 2016). Despite the reasons for this phenomenon, our study supports this trend and highlights its extent in the Argentinian context.

One alarming finding is that almost half of the UDIDs were issued for individuals over 8 years old. This is concerning because ASD symptoms and warning signs can typically be observed during the first few years of life (APA, 2022). This delayed issuance of UDIDs suggests that there may be difficulties in the early detection of ASD from the perspective of both families and healthcare professionals. The difficulties in the detection of ASD in females may also contribute to this problem. Previous studies have shown a significant delay in ASD diagnosis despite early recognition of signs (Montiel-Nava et al., 2023), which is a shared characteristic across high and low-income countries (Matos et al., 2022). Early detection of ASD is crucial for a better prognosis, allowing for therapeutic interventions to take place during developmental periods with more neuroplasticity, while also providing support to the family and community (Freitas & Revoredo, 2022; Molina, 2012; Webb & Jones, 2017).

Typically, the recommendation to issue a UDID comes from the professional who provides the diagnosis. Therefore, the fact that almost half of the UDIDs were issued to individuals older than 8 years old, in a context where most ASD programs are designed for children under the age of 3 (Lopez, 2014; Shrestha et al., 2019), presents a significant challenge.

Regarding the diagnose that justifies the issuance of the UDIDs, PDD is the most common condition, which is a comprehensive diagnosis (Pedreira Massa & González de Dios, 2017) that includes several conditions such as Infantile Autism, Atypical Autism, among others (WHO, 1992). This highlights the current lack of specificity in the diagnosis of ASD. Diagnosing ASD is a complex task that requires consideration of many factors, including developmental age, a spectrum of symptoms, multiple sources of information, and the absence of defined biomarkers (Skellern et al., 2005). Despite these challenges, healthcare professionals must be trained to diagnose autism accurately and precisely, as the diagnosis will determine the therapeutic approach and its outcomes.

### Trends by year in the issuance of UDIDs for autism

Our analysis revealed that male predominance, PPD diagnosis predominance, and delayed issuance of UDIDs were observed globally each year since 2016, indicating stable epidemiological patterns. This underscores the importance of designing targeted interventions to address the persistent issues identified. Although the gender imbalance may be linked to the disorder’s epidemiology, the high prevalence of non-specific diagnoses and delayed processing of UDIDs pose significant health concerns that require further research and actions. Despite recent efforts to promote early diagnosis and train mental health professionals in ASD diagnosis, such as the Argentinian Law number 27.042 enacted in 2014, implementing these policies in real-life scenarios remains a challenge.

Regarding gender, even though the number of UDIDs issued each year is higher for male than for females, in the last two years this difference has narrowed. This increase in women’s UDIDs issuance for autism may reflect better healthcare capacity of identifying the autism female phenotype (Hull et al., 2020; Kirkovski et al., 2013; Loomes et al., 2017).

Although most demographic and diagnosis characteristics remained stable over time, we observed a significant change in the issuance of UDIDs between 2016 and 2021. Specifically, the number of UDIDs issued has decreased since 2020, coinciding with the onset of the Covid pandemic. This may indicate challenges in accessing healthcare services for diagnoses and UDID issuance during this time (Baweja et al., 2022; Musa et al., 2021; Smile, 2020). In fact, studies conducted in Argentina during the pandemic have revealed experiences of exclusion from care and services for autism, exacerbating the risk already reported in normal situations (Valdez et al., 2021).

### Limitations and future directions

The present study has some limitations that should be considered. Firstly, due to the nature of the data, we were unable to perform associative or comparative analyses. Secondly, UDID issuance data is an indirect measure of ASD epidemiology, and not all individuals with ASD diagnoses request a UDID, particularly those requiring minimal support. Additionally, we only had access to data on age, gender and diagnosis. However, previous studies have shown that age and gender are between the most important factors in predicting the occurrence of autism (Hsu et al., 2012). Thus, despite these limitations, our study provides a comprehensive overview of the epidemiology of individuals with ASD in the Autonomous City of Buenos Aires, which was previously unknown and is now supported by official records.

In conclusion, our findings have significant implications for public health by providing a strong epidemiological foundation. On one side, we shed light on the epidemiology of individuals with ASD in the Autonomous City of Buenos Aires, finding that it follows global-wide trends. Therefore, our findings offer valuable insights into the demographics of this population. Future research can build upon our findings and investigate the characteristics of this population at different locations and at national level. On the other side, we identified chronic issues in the diagnosis of ASD, specifically the significant delay and the lack of specificity in diagnosis. These issues should be prioritized on the public agenda to bring attention to the urgent need for improvements in the quality of life of individuals with ASD.

## Data Availability

All data produced in the present work are contained in the manuscript

## Statements and Declarations

The authors have no relevant financial or non-financial interests to disclose.

The authors report there are no competing interests to declare.

